# Detection of SARS-CoV-2 infection by saliva and nasopharyngeal sampling in frontline healthcare workers: an observational cohort study

**DOI:** 10.1101/2021.04.23.21255964

**Authors:** Naomi F. Walker, Rachel L. Byrne, Ashleigh Howard, Elissavet Nikolaou, Madlen Farrar, Sharon Glynn, Katerina S. Cheliotis, Ana I. Cubas Atienzar, Kelly Davies, Jesus Reine, Zalina Rashid-Gardner, Esther L. German, Carla Solórzano, Tess Blandamer, Lisa Hitchins, Christopher Myerscough, Brad Gessner, Elizabeth Biegner, Andrea M. Collins, Mike Beadsworth, Stacy Todd, Helen Hill, Catherine F. Houlihan, Eleni Nastouli, on behalf of the SAFER investigators, Emily R. Adams, Elena Mitsi, Daniela M. Ferreira

## Abstract

**Background:** The SARS-CoV-2 pandemic has caused an unprecedented strain on healthcare systems worldwide, including the UK National Health Service (NHS). During the first wave of SARS-CoV-2 transmission in UK, SARS-CoV-2 NHS diagnostic test availability was limited to self-isolating symptomatic staff. The burden of symptomatic and asymptomatic infection in healthcare workers (HCW) attending work was unknown.

**Methods:** We conducted an observational cohort study of SARS-CoV-2 infection in HCW working in an acute NHS Trust during the first wave of the COVID-19 pandemic, using serial self-collected saliva and nasopharyngeal (NP) samples. We also collected self-assessed symptom profiles and isolation behaviours. We retrospectively compared SARS-CoV-2 detection by RT-PCR from saliva (weekly) and NP swabs (twice weekly) from 85 individuals in this cohort and evaluated the association with symptoms.

**Findings:** Over a 12-week period from 30^th^ March 2020, 40% (n=34/85, CI95% 31.3-51.8%) HCWs had evidence of SARS-CoV-2 infection by surveillance NP swab and/or saliva RT-qPCR. Agreement between paired saliva and NP swabs was poor (28.6%, CI95% 13.2-48.7%) with both methods detecting symptomatic and asymptomatic infections. Symptoms were reported by 47.1% (n=40) and self-isolation by 25.9% participants (n=22). Only 41.2% (n=14/34) participants with SARS-CoV-2 infection reported any symptoms within 14 days of the infection.

**Interpretation:** HCWs are a potential source of SARS-CoV-2 transmission in hospitals and symptom screening will identify the minority of infections in HCW. Saliva is an easily accessible fluid sample for screening for SARS-CoV-2 infection and in addition to NP swab, facilitated ascertainment of symptomatic and asymptomatic cases in this setting. Combined saliva and NP testing would improve detection of SARS-CoV-2 for surveillance. Better understanding of transmissibility from asymptomatic staff using transmission-based infection precautions, is required to inform policy.

## Introduction

The Severe Acute Respiratory Syndrome Coronavirus 2 (SARS-CoV-2), the causative agent of the coronavirus disease 2019 (COVID-19) pandemic, has caused an unprecedented strain on health systems worldwide, including in the United Kingdom National Health Service (NHS) and the frontline healthcare workers (HCW) it employs. HCWs are particularly vulnerable to SARS-CoV-2 infection due to their frequent close proximity to infectious COVID-19 patients (Korth *et al*., 2020). The risk of transmission of SARS-CoV-2 from infected HCW to their patients and colleagues whilst attending work was also recognized as a major threat to infection prevention and control. Transmission-based precautions (including PPE) were implemented widely, for the protection of the staff and their patients.

The global SARS-CoV-2 pandemic response has been hindered by inadequate access to diagnostics. Whilst reverse transcription polymerase chain reaction (RT-qPCR) assays for SARS-CoV-2 on respiratory samples have proven the mainstay of clinical diagnosis, in the early stages of the pandemic, testing capacity was extremely limited, and was initially reserved only for hospitalised individuals meeting the COVID-19 case definition. The UK COVID-19 case definition evolved during the outbreak, losing the initial geographic restrictions on 12^th^ March 2020 as transmission became widespread (see Box 1). Loss of taste and smell was added as a symptom to the case definition in May 2020. With coordinated national and global efforts, diagnostic testing capacity has been scaled up, substantially. Currently, regular asymptomatic screening of HCW is standard practice in the NHS utilizing both respiratory swabbing and saliva, however, there has not been consensus on the optimal frequency or sample type.

### Box 1

**UK COVID-19 Case Definitions**

**Stay at home guidance (for HCW & public)\***

(issued by Chief Medical Officer, 12^th^ March 2020):

a. A new continuous cough OR

b. High temperature (of 37.8 degrees centigrade or higher)

**regardless of travel history or contact with confirmed cases**.

**Current UK government testing advice**

(from https://www.gov.uk/coronavirus, accessed 12^th^ January 2021):

**If you have any coronavirus symptoms:**

- a high temperature
- a new, continuous cough
- a loss of, or change to, your sense of smell or taste**

**Get a test** and stay at home

SARS-CoV-2 Acquisition in Frontline Health Care Workers – Evaluation to Inform Response (SAFER) was a cohort study designed to prospectively collect data on infections in HCW from March – July 2020, in order to inform control strategies in healthcare settings. In this retrospective analysis of a subgroup of SAFER Liverpool participants, we evaluated the prevalence of SARS-CoV-2 RNA detected by weekly saliva or twice weekly nasopharyngeal (NP) self-testing by RT-qPCR, and the occurrence of symptoms of infection. Laboratory analysis was not conducted in real time and HCW followed national guidance for isolation based on symptoms. We and others have previously demonstrated the utility of saliva as a reliable alternative to nasal swabbing for the detection of SARS-CoV-2 in symptomatic individuals (Anne L. Wyllie *et al*., 2020; Byrne *et al*., 2020; Thompson and Cunniffe, 2020; Bastos *et al*., 2021). We compare SARS-CoV-2 detection by RT-qPCR in saliva and nasopharyngeal swabs collected routinely in a mixed cohort of symptomatic and asymptomatic healthcare workers and report the association with symptom profiles.

## Methods

### Ethics

Ethical approval was obtained from the NHS Health Research Authority (ref 20/SC/0147). All participants provided written informed consent.

### Participant recruitment and involvement

Participants were healthcare workers in a variety of roles who enrolled onto the SAFER Study between 30^th^ March and 9^th^ April 2020 at Royal Liverpool University Hospital (RLUH), Liverpool University Hospitals NHS Foundation Trust. Eligible individuals were at least 18 years old and were working in a patient facing role for at least five hours for at least one day during the study period (12 weeks from enrolment). Participants were invited from areas caring for COVID-19 patients (Accident and Emergency (A/E), the acute medical unit, infectious diseases and respiratory wards), and areas which aimed to be COVID-19 free (haematology, surgical wards). Participants were free to withdraw from the study at any point and could choose to either allow their existing data to remain in the study or be excluded. Inclusion in this subgroup analysis required the participant to have provided saliva samples.

### Sample collection

Participants were asked to provide a self-collected NP sample by swabbing the throat and then nose twice weekly when attending work. Swabs were transported in 1ml of liquid amies (MWE, UK) to designated collection points within the hospital by participants and then transported to the laboratory daily by the research team, where they were stored at -80°C, until further analysis. Saliva (at least 500µl) was collected once per week into a sterile tube (SARSTEDT, USA) by passive drool using a plastic funnel at home. Participants were asked to store saliva samples in their home freezers (approximately -20°C) before transportation in cooler packs (provided by the research team) to the Liverpool School of Tropical Medicine (LSTM) laboratories for processing at monthly intervals. Participants were instructed not to collect NP samples during periods of self-isolation, however, they continued to collect weekly saliva samples during this time.

### RNA extraction

Viral RNA from NP swabs was extracted using the high-throughput Quick DNA/RNA™ viral MagBead kit (Zymo, USA), whereas viral RNA from saliva was extracted using the QIAamp Viral RNA Mini Kit (Qiagen, Germany), following in-house optimisation. Both followed manufacturer’s instructions and an internal extraction control incorporated at the lysis stage (Genesig, UK). Once extracted, samples were taken immediately for downstream application and stored on ice during RT-qPCR setup.

### SARS-CoV-2 RT-qPCR

For SARS-CoV-2 RT-qPCR detection, 8µl of extracted RNA was tested using the Genesig® Real-Time Coronavirus COVID-19 PCR assay (Genesig, UK) in a RGQ 6000 thermocycler (Qiagen, Germany). Samples were classified as RT-qPCR positive if both the internal extraction and the SARS-CoV-2 probes were detected at <40_Ct_. Genome copy numbers per ml (gcn/ml) were quantified using the manufacturer’s positive control (1.67 × 10^5^ gcn/µl) as a reference for threshold. Samples with invalid RT-qPCR internal extraction amplification results were re-extracted and re-run.

### Analytical sensitivity

The analytical sensitivity of saliva compared to NP swabs was compared by spiking the samples with serial dilutions of SARS-CoV-2 culture supernatant. The isolate REMRQ0001/human/2020/Liverpool propagated in VERO E6 cells and maintained as previously described (Patterson *et al*., 2020) was used for the serial dilutions. Fourteen NP swabs were stored in 1ml of amies preservation medium (Copan, Italy) and 4ml of saliva were self-collected from a confirmed RT-qPCR SARS-CoV-2 negative volunteer. A serial dilution series of SARS-CoV-2 ranging from 10^6^ to 10^−6^ plaque forming units per ml (pfu/ml) was used to spike 140µl of saliva and swab samples. The limit of detection (LOD) was determined by the lowest concentration for which all three RT-qPCR replicates amplified. For quantification of the gcn/ml, viral RNA of the serial dilutions was extracted using QIAmp Viral RNA mini kit (Qiagen, Germany) and the gcn/ml were calculated using the COVID-19 Genesig RT-qPCR kit (PrimerDesign, UK) with a ten-fold serial dilution of quantified specific in vitro-transcribed RNA (Genesig, UK).

### Symptom reporting, isolation and diagnostic tests

Symptom reporting was via a short questionnaire that was completed twice weekly, accompanying each sampling episode; participants could select from COVID-19 case definition symptoms and other acute respiratory illness symptoms or enter other symptoms in free text. At the time of the study, SARS-CoV-2 testing of staff who did not meet the UK COVID-19 case definition (Box 1) was not routine, nor feasible due to lack of laboratory capacity and testing reagents. Participants with symptoms that met the COVID-19 case definition were advised to self-isolate and seek COVID-19 diagnostic testing via the staff testing service when it became available but also continued study procedures, except for self NP collection during periods of self-isolation. Periods of self-isolation and standard-of-care test results were reported by participants to the research team and recorded. Symptoms associated with each sample collected were scored on a scale from 0 (no symptoms) to 6 (multiple symptoms consistent with the COVID-19 case definition) for the purposes of analysis (see Box 2). Participants were assigned the highest code for which they qualified.

Study samples were stored until the end of the study follow up period, and laboratory testing for SARS-CoV-2 infection was retrospective. Results were made available to participants after the laboratory analysis was complete.

#### Box 2

**Symptom scores assigned for symptom and isolation status**

| **Box 2 Symptom scores assigned for symptom and isolation status** |  |
| --- | --- |
| **0** | Asymptomatic |
| **1** | Symptom (s), likely unrelated e.g. ankle pain |
| **2** | Symptom (s), non-specific e.g. Fatigue / headache / muscle aches |
| **3** | One respiratory symptom, not COVID case definition<br>e.g. Cough (other), sore throat, coryza |
| **4** | More than one symptom including a respiratory symptom<br>(not COVID-19 case definition symptom) |
| **5** | COVID-19 case definition symptom: New/continuous cough or fever or loss of taste/smell |
| **6** | More than one COVID-19 case definition symptom: new/continuous cough, fever, loss of taste or smell |

### Statistical analysis

Analysis was in R (version 4.0.4) and Prism 8 (Graphpad). Two-tailed statistical tests was used to compare variables throughout the study. Non-parametric Mann-Whitney test was used to compare two groups. Correlations were assessed using Spearman’s correlation test using raw, non log-transformed values.

## Results

### Participants and follow up

In total, 99 participants were enrolled into the SAFER Liverpool cohort. Of these, 85 participants provided both saliva and NP samples over 8 and 12 weeks respectively and are therefore included in this analysis. Participant characteristics are reported in Table 1. The majority (77.6%, n=66) were female. Participants comprised doctors, nurses, healthcare assistants and allied health professionals (physiotherapists and occupational therapists). Retention in follow up was 84.7%, (n=72) participants at four weeks, 81.2% (n=69) participants at eight weeks and 74.1% (n=63) (74.1%) participants at twelve weeks. A median of 21 NP swabs (interquartile range (IQR) 10.5-24.0) of a target of 24 and 8 saliva samples (IQR 4.50-8.00) of a target of 8 were submitted per participant.

**Table 1.** Participant characteristics.

|  |  |
| --- | --- |
| Total included participants (n) | 85 |
| Female, n (%) | 66 (77.6) |
| Age, median years (IQR) | 35.0 (27.5-46.5) |
| Role, n (%) |  |
| Doctor | 35 (41.2) |
| Nurse | 32 (37.6) |
| Health Care Assistants | 9 (10.6) |
| Allied health professional | 6 (7.1) |
| Other | 3 (3.5) |
| Experienced symptoms*, n (%) |  |
| Any | 40 (47.1) |
| COVID-19 case definition | 9 (10.6) |
| Isolated during study period, n (%) | 22 (25.8) |
| COVID-19 diagnosis during study** | 3 |
| COVID-19 hospitalisation | 0 |

\*Over 12 weeks study period
\*\*outside of the study setting

### COVID-19 diagnosis, symptoms and isolation status

Of the 85 participants, 40.0% (n=34, CI95% 31.3-51.8%) were positive for SARS-CoV-2 RNA on at least one study sample during the study period. Symptoms of any kind were reported by 47.1% (n=40) participants on twice weekly questionnaires during study follow up whilst attending work. However, only 41.2% (n=14/34) participants with SARS-CoV-2 RNA detected by SAFER study sampling reported symptoms within a 14-day period either side of their positive result (Table 2). In participants with evidence of SARS-CoV-2 infection attending work, symptoms were predominantly respiratory (coryza, sore throat, cough) but most often not those compatible with the COVID-19 case definition.

**Table 2.** Symptoms reporting in SARS-CoV-2 RNA positive participants during study follow up. (excluding participants who were self-isolating for COVID-19 case definition symptoms)

|  |  |
| --- | --- |
| Participants with positive results (NP swab or saliva), n | 34 |
| Participants with symptoms associated with positive result*,<br>n (% positive participants) | 14 (41.2) |
| Fever | 0 (0) |
| New continuous cough | 0 (0) |
| Cough, other | 6 (23.6) |
| Rhinorrhoea | 8 (23.5) |
| Muscle aches | 1 (2.9) |
| Joint Pain | 2 (5.9) |
| Sore Throat | 5 (14.7) |
| Headache | 4 (11.8) |
| Chest Pain | 0 |
| Breathing Difficulty | 0 |
| Change in Taste or Smell** | 5 (14.7) |

\*within 14 days of positive result
\*\*at any time during study follow up

The timing of SARS-CoV-2 RNA detection and relationship with symptoms and self-isolation is shown in Figure 1. Most positive results were detected in the early weeks of the study period, which commenced shortly after the first national lockdown was declared (23^rd^ March 2020) and prior to universal use of face masks/coverings at all time for staff in hospital (introduced on 15^th^ June 2020). Nine participants reported symptoms consistent with the COVID-19 case definition and 14 participants reported COVID-19 testing during the 12-week period, of which only 3 participants received a confirmed diagnosis of COVID-19 by qPCR of a nose/throat sample, for clinical purposes outside of the study setting. Periods of self-isolation were prevalent, reported by 25.9% (n=22/85) participants. In 7 incidences, self-isolation was reportedly due to a contact rather than due to symptoms. Eight participants who self-isolated tested positive for SARS-CoV-2 RNA on study samples. Only 10/34 participants who were SARS-CoV-2 RNA positive self-isolated at any point during the study. The median duration of isolation per participant was 6.5 (IQR 5.0-14.0) days.

**Figure 1.**
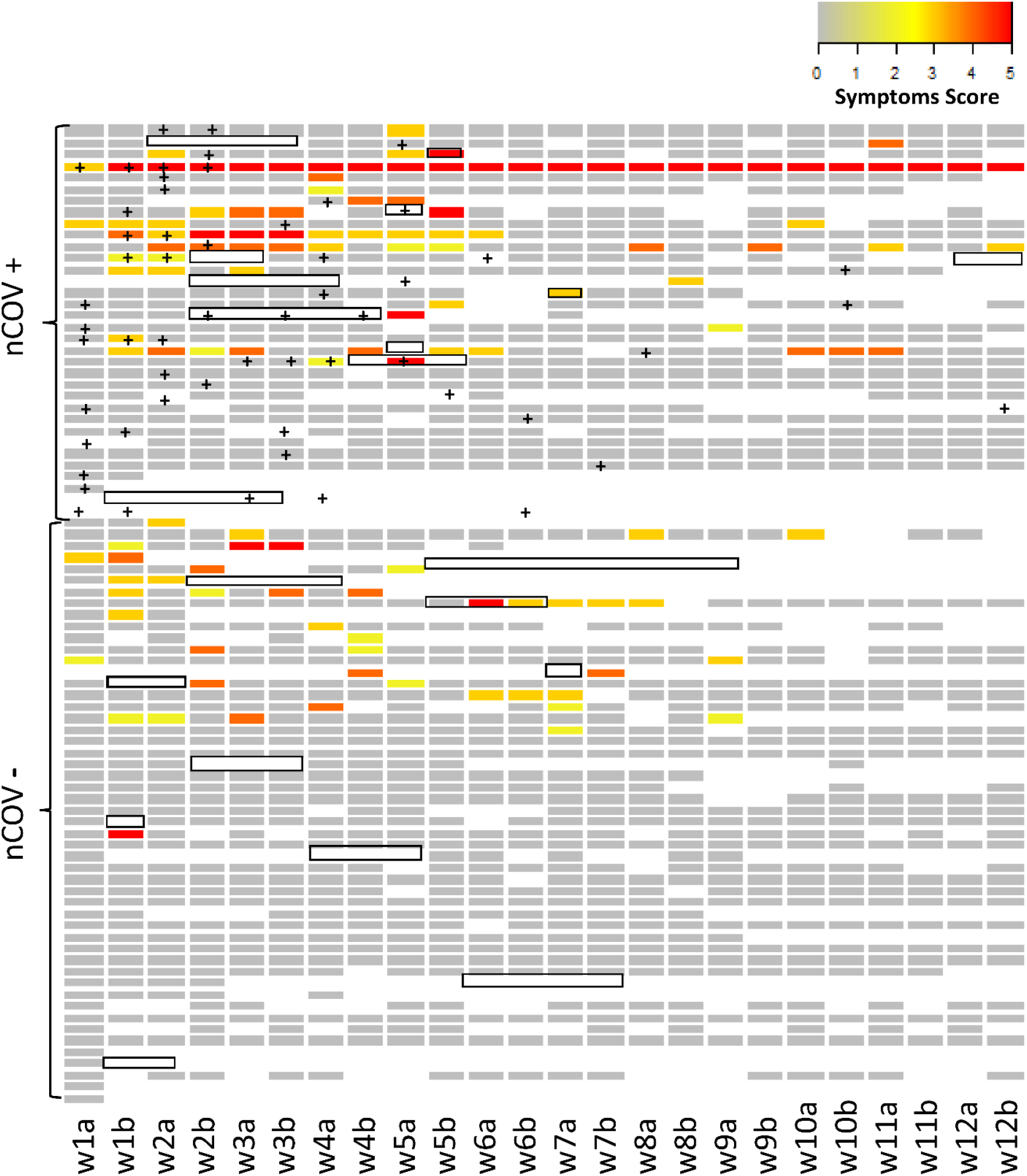
Symptoms, isolation and SARS-CoV-2 RNA status in HCW (n=85) over a period of 12 weeks. Heatmap for recorded symptoms using a colour scale to represent symptoms score (see Box 2) from grey (0: Asymptomatic) to red (5: a symptom consistent with the COVID-19 case definition in red). No participants scored 6. The participant with symptoms recorded at every timepoint experienced loss of smell which persisted throughout the study period. Isolation period is by a black box delineating timepoints. Participants were clustered into two groups, nCoV+ (positive for SARS-CoV-2 RNA by either diagnostic method), or nCoV- (SARS-CoV-2 RNA negative on results available). Time points with missing data are shown in white.

### Analytical sensitivity

The analytical sensitivity in spiked samples indicated the LOD for saliva was 10^−2^ pfu/ml ^(^ ≈ 2.0 × 10^1^ gcn/ml) and for the NP swabs 10^0^ pfu/ml ^(^ ≈ 2.0 × 10^3^ gcn/ml). However, amplification of one or more replicates was recorded for saliva and NP swabs to concentrations 10^−6^ (pfu/ml) and 10^−4^ (pfu/ml), respectively.

### Comparison of saliva and NP swab for detection of SARS-CoV-2 infection by qPCR

SARS-CoV-2 RNA was detected in 3.02% (n=43/1425) NP samples and 4.15% (n=22/530) saliva samples (Figure 2). Overall, 8 participants were positive for SARS-CoV-2 RNA on both NP swab and saliva, 17 participants were positive by NP swab only and 9 participants by saliva only. Reports of symptoms within 14 days of a positive result were similar in participants who were positive by NP swab compared to saliva (44.0%, (n=11/25) and 40.0% (n=8/16) respectively, p=0.757). The median viral load in SARS-CoV-2 RNA positive participants with symptoms was 139 gcn/ml- (IQR: 31-3332 gcn/ml) and without symptoms was 64.4 gcn/ml (IQR: 25.1-494.5 gcn/ml), p=0.65. No correlation was found between viral loads and assigned symptom score (r=-0.12, p= 0.52).

**Figure 2.**
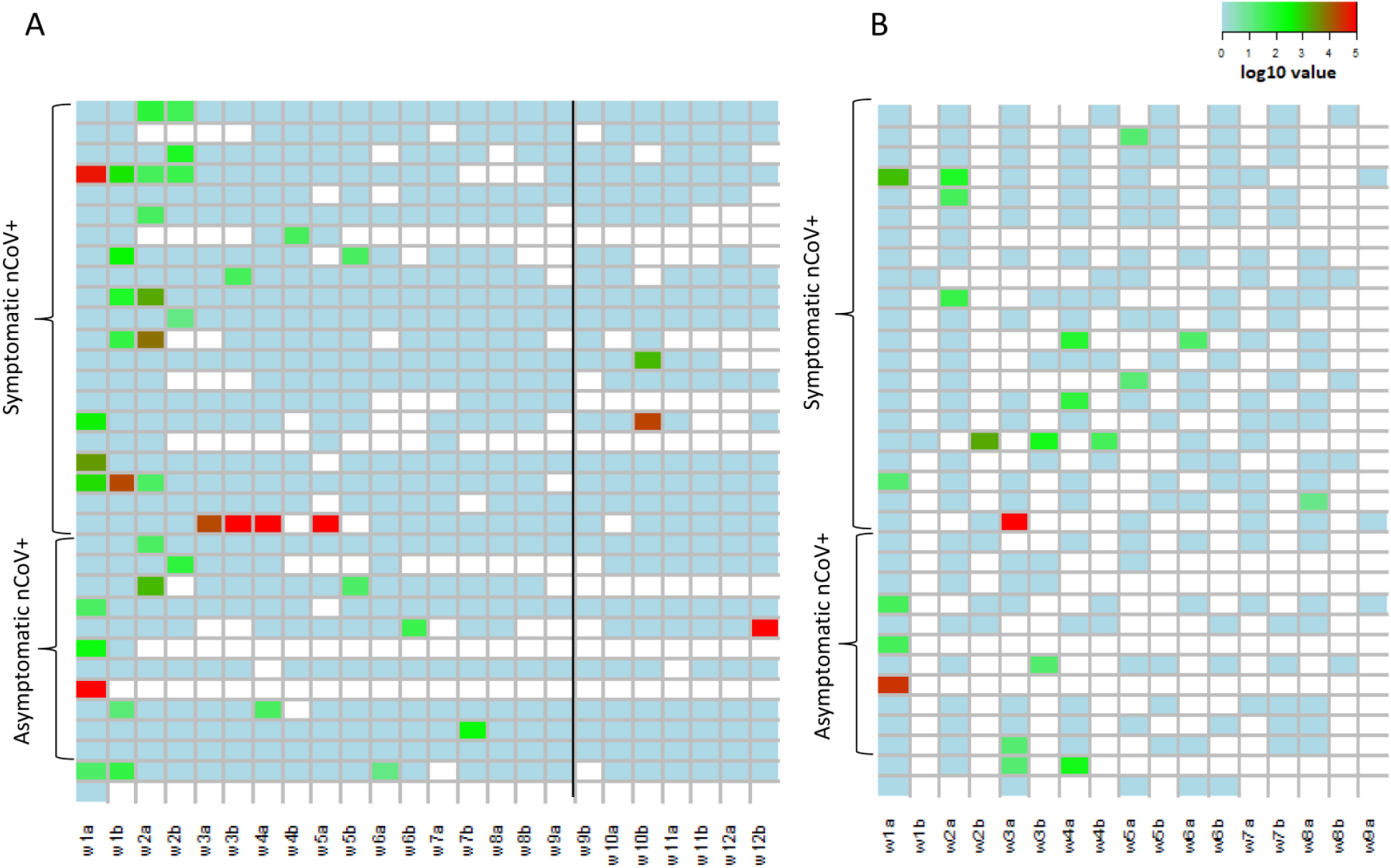
SARS-CoV-2 RNA detection by saliva and nasopharyngeal (NP) swab during study period. Timing of SARS-CoV-2 positivity by NP swab and saliva sample over a period of 12- and 8-weeks, respectively, in SARS-COV-2 RNA positive (nCoV+) participants, clustered by symptom status (symptomatic or asymptomatic). **A)** Heatmap of log10 viral load value per NP swab sample over surveillance period of 12 weeks (n= 25 nCoV+ by NP swab, 15 symptomatic and 10 asymptomatic). **B)** Heatmap of log10 viral load value per saliva sample over the sampling period of 8 weeks (n=17 nCoV+, 11 symptomatic and 6 asymptomatic).

### The agreement of saliva and NP swabs for SARS-CoV-2 detection in a paired sample cohort

We then investigated the agreement of NP swabs and saliva for SARS-CoV-2 detection, utilizing paired samples where both saliva and NP samples had been self collected on the same date. A total of 225 paired samples were collected, of which 20 (8.89%) included a SARS-CoV-2 RNA positive result. This comprised 12 positive NP samples and 13 positive saliva samples. Of these, 7 were positive exclusively by NP swab and 8 exclusively by saliva. In only 5 paired samples were the NP and saliva result both positive. There was no significant difference between viral loads for either specimen (p=0.312) as shown in figure 3. For SARS-CoV-2 RNA positive individuals with paired samples, this resulted in 35.0% (n=7/20) positive only by NP swab, 40.0% (n=8/20) positive only by saliva and 25.0% (n=5/20) consistently positive for both specimens. This produced an estimated prevalence in this paired sample subgroup of 73.5% and 50.0% for NP swabs and saliva respectively, comprising samples from 83 individuals. The proportion positive agreement of saliva and NP swabs was 40.0% with an overall agreement of 93.3%.

**Figure 3.**
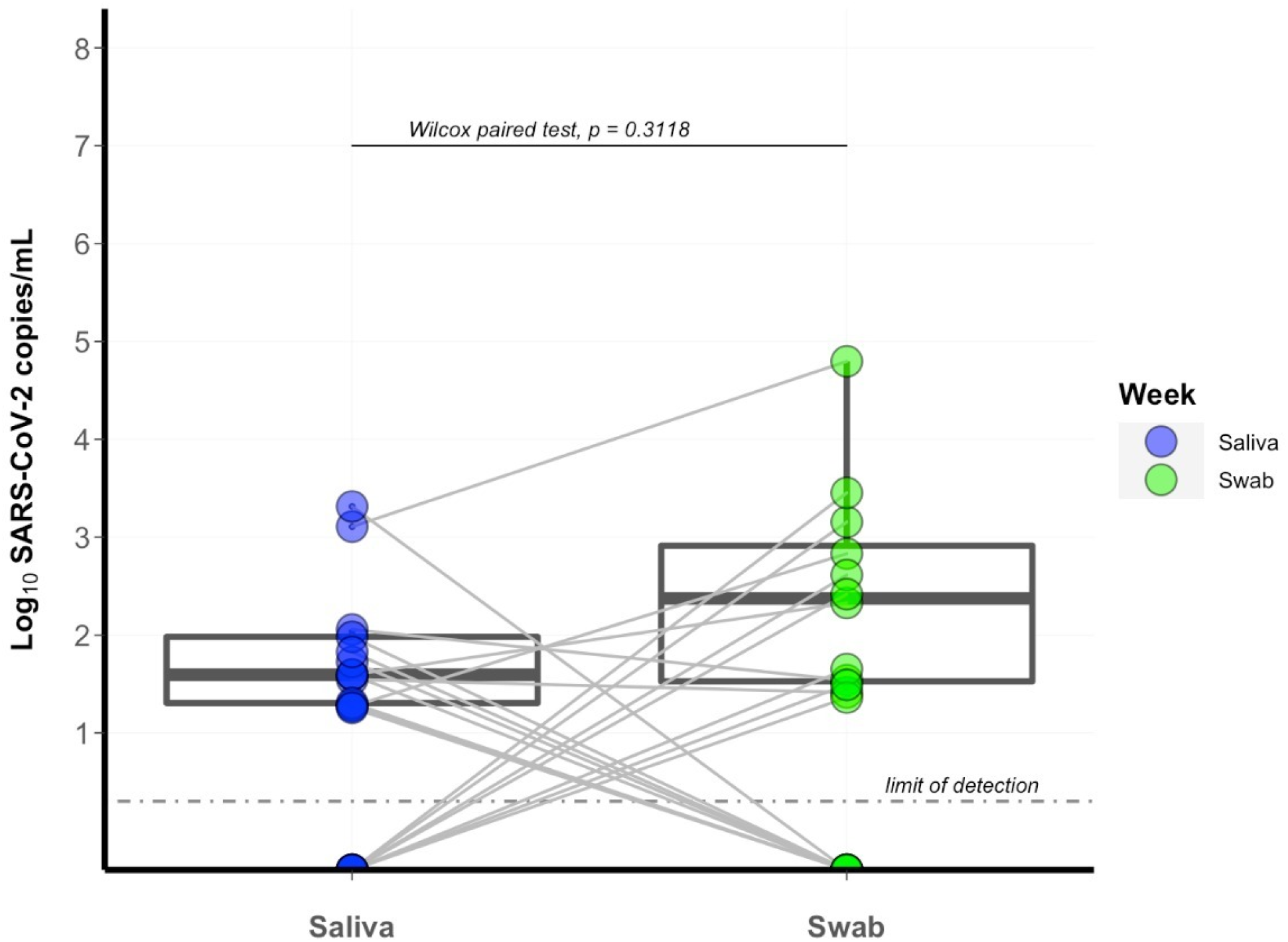
Comparison of SARS-CoV-2 viral load in paired nasopharyngeal swab and saliva samples. The logged viral load of paired saliva and nasopharyngeal (NP) swabs (n=20) from healthcare workers for the first 8 weeks of the study period is shown. Paired samples were those where the participant self-collected both saliva and NP swab on the same date. Data points for a single sample are represented by a point outlined by a circle and a single line connects paired samples. Boxes represent interquartile range (IQR) with a median central line.

## Discussion

Here, we describe a cohort of 85 frontline HCW in Liverpool and present results of surveillance NP swabs and saliva samples for SARS-CoV-2 infection, for up to 12 weeks during the first peak of COVID-19 pandemic. A high proportion (40%) of frontline healthcare workers had evidence of SARS-CoV-2 infection during study follow up. Most infections occurred early in the follow up period, likely reflecting transmission prior to community interventions and widespread use of transmission-based precautions in hospital. This supports findings from other UK healthcare settings demonstrating a high prevalence of SARS-CoV-2 infection in HCW in the NHS in Spring 2020 (Houlihan *et al*., 2020;Keeley *et al*., 2020; Treibel *et al*., 2020).

We report poor agreement between saliva and NP swabs for the detection of SARS-CoV-2. This is consistent with reports by Rao *et al*., (2020) in asymptomatic adult males detained in a COVID detention centre, where exposure to SARS-CoV-2 is presumed to be high. Most other data pertaining to the performance of saliva for SARS-CoV-2 diagnosis is derived from symptomatic individuals, with numerous studies reporting good concordance (Wyllie *et al*., 2020a, 2020b; Byrne *et al*., 2020), as also reported by Lee *et al*. (2020), in a recent systematic review combining 37 studies of which the majority of participants were symptomatic, where a small difference between the sensitivity of saliva and NP swabs (−3.4% (95% CI -9.9 to 2.1%) was found. Interestingly, here we report from spiked saliva samples, that SARS-CoV-2 can be detected with greater sensitivity in saliva compared to NP swabs by 100-fold As we continue to see SARS-CoV-2 cases reduce it will be important to quickly and accurately detect asymptomatic cases in the community. It is important to note that in our paired analysis, 26% (n=9) SARS-CoV-2 infections were missed by NP swabs and thereby saliva should still be considered an alternative to more invasive, more costly swabbing (Bastos *et al*., 2021). This study supports the use of saliva in addition to NP sampling for optimal detection of infection.

This dataset is unique in including both regular saliva analysis and NP swabbing into a HCW surveillance study. As routine asymptomatic staff testing was not available during the study period and laboratory capacity at the time did not permit contemporaneous analysis, the study results were not available until after the study period and therefore did not influence participant symptom reporting or self-isolation behavior. A minority of symptomatic participants with SARS-CoV-2 infection met the COVID-19 case definition. A large proportion (58.8%) of SARS-CoV-2 infections detected in this HCW cohort were asymptomatic. This is considerably higher than predicted from a recent systematic review and meta-analysis which found that the minority of infections were asymptomatic (estimated 20%, 95% confidence interval 17-25) (Buitrago-Garcia *et al*., 2020).

Regular screening of HCW, irrespective of symptoms has now been implemented in UK as an intervention to control SARS-CoV-2 transmission in hospitals and other healthcare settings. However, the impact of this policy in terms of staff absences and the potential indirect consequences for the healthcare system has not yet been realised. Transmission from asymptomatic individuals may be low, especially if these individuals are trained in transmission-based precautions and are using appropriate PPE (Buitrago-Garcia *et al*., 2020; Madewell *et al*., 2020). Poor staffing levels in acute settings may pose a risk to patients and may increase the psychological burden on staff. It is essential to gain further data on the transmissibility of asymptomatic infection in this context to fully appreciate the risk-benefit of intensive HCW surveillance interventions (Gandhi, Yokoe and Havlir, 2020; Pollock and Lancaster, 2020).

Finally, we note that HCW did not consistently self-isolate when respiratory symptoms were experienced. Resource limitations reducing access to routine diagnostic testing possibly impacted on staff isolation and test-seeking behaviour in the first wave of the UK COVID-19 pandemic. Occupational testing first became available to symptomatic HCW at Liverpool University Hospitals NHS Foundation Trust on 2^nd^ April 2020. However, presenteeism in HCW may be a contributory factor, even in the context of a global respiratory pandemic (Tartari *et al*., 2020). Clarification on and reinforcement of self-isolation and testing policies for HCW when symptomatic is advisable, especially given symptomatic infections are likely more transmissible (Buitrago-Garcia *et al*., 2020).

Limitations of this study include some loss to follow up and withdrawal of participants with resulting missing data. However, the overall retention in the study was good (over 80% at week 8) and return of samples was overall high. There was possible inconsistency of self-sampling technique as participants were not supervised. Sample processing, which required one freeze-thaw cycle (NP swab) or two freeze-thaw cycles (saliva) prior to RNA extraction, possibly reduced the overall yield of the PCR assay.

In summary, we report a high prevalence of SARS-CoV-2 infection in asymptomatic and symptomatic HCWs, attending work during the first wave of the COVID-19 pandemic in UK, as detected by saliva and NP surveillance sampling. However, there was limited agreement between the two sampling methods. Detection of SARS-CoV-2 by surveillance sampling far exceeded case detection by routine diagnostic testing (targeted to those with symptoms) and the majority of infections were asymptomatic. This study supports the use of saliva as an additional sample for screening for SARS-CoV-2 infection, in order to optimize case detection of symptomatic and asymptomatic infections. Saliva PCR should therefore be considered in addition to NP PCR for SARS-CoV-2 detection, where high sensitivity screening is required.

## Data Availability

Data available upon request.

## Acknowledgements

We acknowledge and thank the participants of the SARS-CoV-2 Acquisition in Frontline Health Care Workers – Evaluation to Inform Response. We also acknowledge the valuable contributions of LSTM Accelerator Research Clinic staff and volunteers, and LUHFT Clinical Research Unit team who supported the follow up of participants. We thank Natalie Tate and Sherin Pojar for excellent technical and logistical support and Saigini Vickneswaran and Chiamaka Sophie Njoku for support with data management. We thank Dr Ghaith Aljayyoussi for assistance with R code. We acknowledge and thank University College London Hospitals NHS Foundation Trust for sponsorship of the study. N.F.W was funded by the National Institute for Health Research Health Protection Research Unit (NIHR HPRU) in Emerging and Zoonotic Infections, the Centre of Excellence in Infectious Diseases Research (CEIDR) and the Alder Hey Charity. D.F and E.M received funds from Pfizer grant no. WI255862, for this study. We also acknowledge support of Liverpool Health Partners and the Liverpool-Malawi-COVID-19 Consortium.

SAFER Investigators: Jude Heaney, Matt Byott, Dan Frampton,Moira Spyer, Sarah Edwards, Andrew Hayward, Richard Gilson, Ed Manley, Susan Michie, Catherine Houlihan, Eleni Nastouli

## Footnotes

* initially for a minimum of 7 days as no community or occupational testing was available

** introduced into the case definition from 18^th^ May 2020

## Notes

### Competing Interest Statement

The authors have declared no competing interest.

### Clinical Trial

20/SC/0147

### Funding Statement

Liverpool COVID-19 Partnership Strategic Research Fund to NFW; Pfizer award to DF and EM (grant number WI2558862-1). The Medical Research Council doctoral training partnership fund to RLB. CFH and EN received funding from MRC/UKRI (SAFER study; grant number MC PC 19082)

### Author Declarations

NHS health research authority (ref 20/SC/0147)

